# Inspecting Consistency Between US and European Vitamin D Guidelines Using Physiologically-Based Pharmacokinetics Modelling

**DOI:** 10.1101/2025.02.13.25322205

**Authors:** You Tao

## Abstract

According to the US official guideline for vitamin D, serum 25-hydroxyvitamin D over 125 nmol/L is linked to potential toxicity. Using a physiologically-based pharmacokinetics model based on a randomised controlled trial in Cape Town, South Africa, we showed the 2000 IU daily dose, recommended by European Food Safety Agency as a safe dose, is expected to lead to serum concentration exceeding the 125 nmol/L threshold among some children and adolescents. This highlights inconsistency between different guidelines and the need to use modelling to bridge the gap between dose and pharmacokinetics.

## Introduction

The serum levels of vitamin D metabolite 25-hydroxyvitamin D (25(OH)D) is widely accepted as a marker for vitamin D status. In the US, according to the National Academies of Sciences, Engineering, and Medicine (NASEM) and the Endocrine Society, the working definition includes deficiency (< 30 nmol/L), inadequacy (30 – 50 nmol/L), adequacy (50 – 125 nmol/L) and potential toxicity (>125 nmol/L) [1-3].

For children (1–11 years) and adolescents (12–18 years), the Endocrine Society suggests empiric vitamin D supplementation to prevent nutritional rickets and potentially lower the risk of respiratory tract infections [2]. Daily doses between 300 – 2000 IU are assessed in studies on respiratory tract infection prevention, but no specific dose is recommended by the Endocrine Society [2].

According to European Food Safety Agency (EFSA), daily dose up to 2000 IU is safe for children aged 1-10 years [4]. As a first step, a useful goal is to inspect consistency between the NASEM and EFSA guidelines through pharmacokinetics (PK) modelling, which integrates accessible information in an unbiased manner.

So far, PK modelling of orally administered vitamin D has been reported for children with chronic kidney disease [5] and children with obesity and asthma [6]. These reports highlight the importance of a weight-based approach to dose selection. Unfortunately, no modelling is established for healthy children. We developed a Physiologically-Based Pharmacokinetics (PBPK) model based on a 3-year study of healthy schoolchildren in Cape Town, South Africa [7] by customising our previous modelling of healthy adults [8, 9]. Sex and weight were used as covariates to predict the volume of different compartments, and age-normalised body mass index (ZBMI) was used to predict the fat mass. The development and qualification of this model is reported in a different paper [10].

To inspect consistency, our objective is to evaluate how serum 25(OH)D might change under various daily doses in children (6-10 years) and teenagers (11-17 years). To produce a quick answer, we simulated the Cape Town model. Specifically, we simulated 3 safe daily doses for 6- to 17-year-olds, namely 500, 1000 and 2000 IU for 1 year in deficient (10 nmol/L baseline) and adequate cases (50 nmol/L baseline). For the rest of the article, doses are administered daily. Indeed, we identified potential inconsistency between EFSA and NASEM guidelines. Despite the limitations, these simulation results are worth discussing.

## Methods

At each age, 1250 boys and 1250 girls were simulated. For children 6 to 10, body weight was directly sampled according to the WHO report [11]. For adolescents 11 to 17, height [12] and body mass index (BMI) [13] were sampled independently according to the WHO reports to calculate the weight for each person. Weight increase within a year was sampled uniformly for all children and adolescents, respectively. Random effect of the partition coefficient (i.e. ratio between 25(OH)D concentration in the rest of the body and serum) was sampled.

In each simulated study, random effects and residual errors of the model were sampled for each subject, therefore 2500 times, following the distribution inferred from the Cape Town study. At each age, 30 virtual studies were generated to calculate the 2.5^th^ , 50^th^ and 97.5^th^ percentiles, together with 95% prediction intervals around each percentile. Specifically, only additive residuals (mean: 0.0025 nmol/L) but not the proportional residuals (mean: 0.11) were sampled [10] to ensure all subjects started around the same baseline in each simulation.

## Results

We simulated 1-year continuous dosing at daily doses of 500, 1000 and 2000 IU. The 95% prediction intervals for the 2.5^th^ and 97.5^th^ percentiles are small (supplementary Figure S1). Hence, we should focus on the expected values of the percentiles. For clarity, the mean of the 2.5^th^ (Table 1) and 97.5^th^ percentiles (Table 2) of the final values with respect to starting age and baseline are shown. Percentiles under the same dose decrease with age in general. This is intuitive, as older age is associated with higher weight and larger volume of distribution.

**Table 1.** The mean of the predicted 2.5^th^ percentiles after 1-year dosing. Red: <30 nmol/L; yellow: 30–50 nmol/L; green: 50 – 125 nmol/L. Age refers to the starting age.

| Age | 10 nmol/L Baseline |  |  | 50 nmol/L Baseline |  |  |
| --- | --- | --- | --- | --- | --- | --- |
|  | 500 IU | 1000 IU | 2000 IU | 500 IU | 1000 IU | 2000 IU |
| 6 | 31 | 55 | 80 | 62 | 75 | 79 |
| 8 | 29 | 47 | 76 | 60 | 70 | 80 |
| 9 | 26 | 44 | 74 | 60 | 70 | 80 |
| 10 | 25 | 40 | 70 | 60 | 68 | 80 |
| 11 | 25 | 40 | 68 | 61 | 70 | 75 |
| 12 | 24 | 37 | 65 | 60 | 67 | 76 |
| 14 | 20 | 31 | 52 | 58 | 60 | 75 |
| 17 | 19 | 28 | 45 | 57 | 60 | 72 |

**Table 2.** The mean of the simulated 97.5 ^th^ percentiles after 1-year dosing. Grey: >125 nmol/L. Age refers to the starting age.

|  | 10 nmol/L Baseline |  |  | 50 nmol/L Baseline |  |  |
| --- | --- | --- | --- | --- | --- | --- |
| Age | 500 IU | 1000 IU | 2000 IU | 500 IU | 1000 IU | 2000 IU |
| 6 | 75 | 95 | 145 | 80 | 105 | 158 |
| 8 | 65 | 87 | 130 | 76 | 98 | 142 |
| 9 | 63 | 85 | 123 | 75 | 95 | 135 |
| 10 | 60 | 82 | 116 | 74 | 92 | 129 |
| 11 | 60 | 81 | 115 | 75 | 92 | 128 |
| 12 | 58 | 79 | 110 | 74 | 90 | 122 |
| 14 | 50 | 71 | 98 | 65 | 85 | 105 |
| 17 | 45 | 66 | 91 | 68 | 81 | 115 |

For 8–17 years with the 10 nmol/L baseline, the 2.5^th^ percentile was expected to be deficient under the 500 IU dose (red in Table 1). For 8–17 years under 1000 IU, the 2.5^th^ percentile was expected to be inadequate (yellow in Table 1). This is expected, as severely deficient cases with 10 nmol/L baseline typically need higher loading doses (age 6 months– 11 years: 6000 IU daily; 12–18 years: 10000 IU daily) for 8–12 weeks [14].

For almost all other conditions, the 2.5^th^ percentile was expected to be adequate after a year (green in Table 1). Interestingly, for cases with 50 nmol/L baseline, both 2.5^th^ and 97.5^th^ percentiles of the 1000 IU group are expected to remain adequate for 6-11-year-olds.

The 97.5^th^ percentiles were expected to exceed the 125 nmol/L threshold for 6-8-year-olds with 10 nmol/L baseline and 6-11-year-olds with 50 nmol/L baseline (grey in Table 2). This is inconsistent with the EFSA guidelines which recommend 2000 IU daily dose as a safe dose. This is discussed next.

In addition, for 12-17 years, the 97.5^th^ percentiles under 2000 IU are expected to be <125 nmol/L (Table 2).

## Discussion

To derive the safe dose for children, EFSA first identified the lowest observed adverse effect level (LOAEL) from two randomised controlled trials, which was 10000 IU/day [4]. The EFSA applied a 2.5-fold uncertainty factor to obtain the upper limit (UL) of a safe daily dose of 4000 IU for anyone over 11 [4]. For children aged 1-10 years, the ESFA daily UL is 2000 IU [4].

In contrast, the 125 nmol/L threshold was mentioned in the NASEM 2011 report [1]. However, throughout the entire report, they did not present any evidence to support the 125 nmol/L threshold. Hence, we place less emphasis on the 125 nmol/L threshold.

Using modelling, we identified inconsistency between EFSA and NASEM guidelines. The only implicit assumption we made was the Cape Town children would follow the same weight distribution reported by the WHO. We checked the Z-score for BMI distribution of the Cape

Town children from the 3-year study [7] and found they indeed followed the same distribution (data not shown for brevity). Hence, the inconsistency might be real.

This result highlights the need for more modelling work to consistently integrate all accessible vitamin D data, including dosage, 25(OH)D levels, with demographic information (at least sex, age and weight). Ultimately, the goal is to inform selection of a dosing strategy to deliver therapeutic benefits or successful disease prevention, which would also require a good understanding of the pharmacodynamics. The PK/PD modelling process is widely accepted for drug discovery and development. Yet, it needs to be adopted for vitamin D research, which traditionally stemmed from the nutritional science area.

## Conclusions

In summary, our modelling work uncovered potential inconsistency between ESFA and NASEM guidelines, which warrants more modelling work to reconcile such differences. This questions the validity of the claim 125 nmol/L should mark toxicity due to the lack of evidence.

## Data Availability

The model described in this paper is available upon request. Academic use is free. It should not be used for any commercial activity without the written approval of the author.

