## Supplementary Information for "Inspecting Consistency Between US and European Vitamin D Guidelines Using Physiologically-Based Pharmacokinetics Modelling"

YOU Tao, PhD[1,2,3]

[1]Beyond Consulting Ltd. 14 Tytherington Park Road, Macclesfield, Cheshire, UK SK10 2EL
[2]Department of Pharmacology & Therapeutics, University of Liverpool, Liverpool, UK L69 3GE
[3]Centre of Excellence for Long-acting Therapeutics (CELT), University of Liverpool, Liverpool, UK L69 3GE

### Supplementary Materials

**Figure S1.** Simulation of serum 25(OH)D concentration for 1-year continuous daily dosing in European children aged 6 (A: 10 nmol/L baseline; B: 50 nmol/L baseline), 8 (C: 10 nmol/L baseline; D: 50 nmol/L baseline), 9 (E: 10 nmol/L baseline; F: 50 nmol/L baseline), 10 (G: 10 nmol/L baseline; H: 50 nmol/L baseline) and adolescents aged 11 (I: 10 nmol/L baseline; J: 50 nmol/L baseline), 12 (K: 10 nmol/L baseline; L: 50 nmol/L baseline), 14 (M: 10 nmol/L baseline; N: 50 nmol/L baseline) and 17 (O: 10 nmol/L baseline; P: 50 nmol/L baseline). The three lines mark the 2.5th, 50th and 97.5th percentiles with shaded areas showing the 95% confidence intervals.

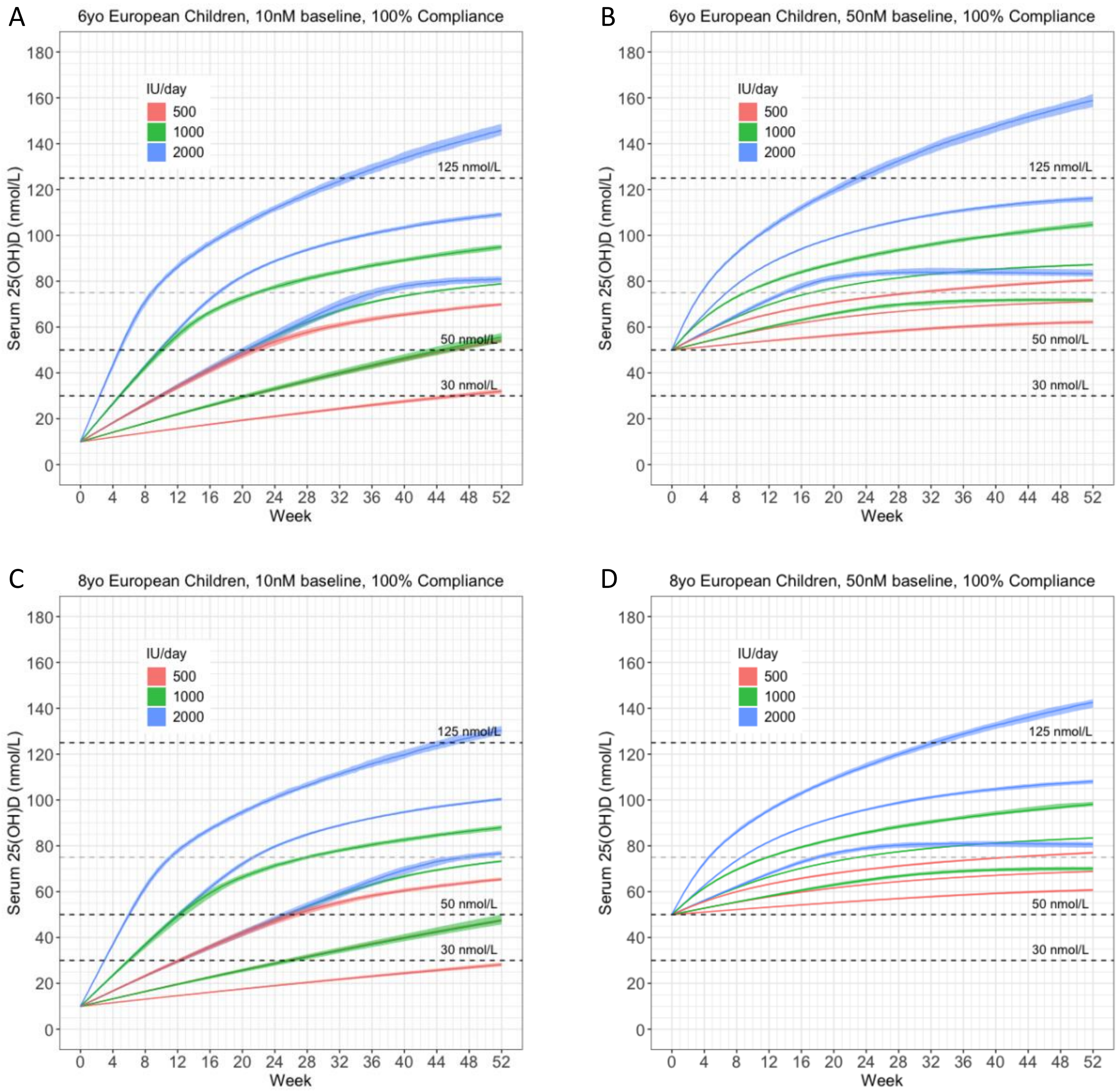

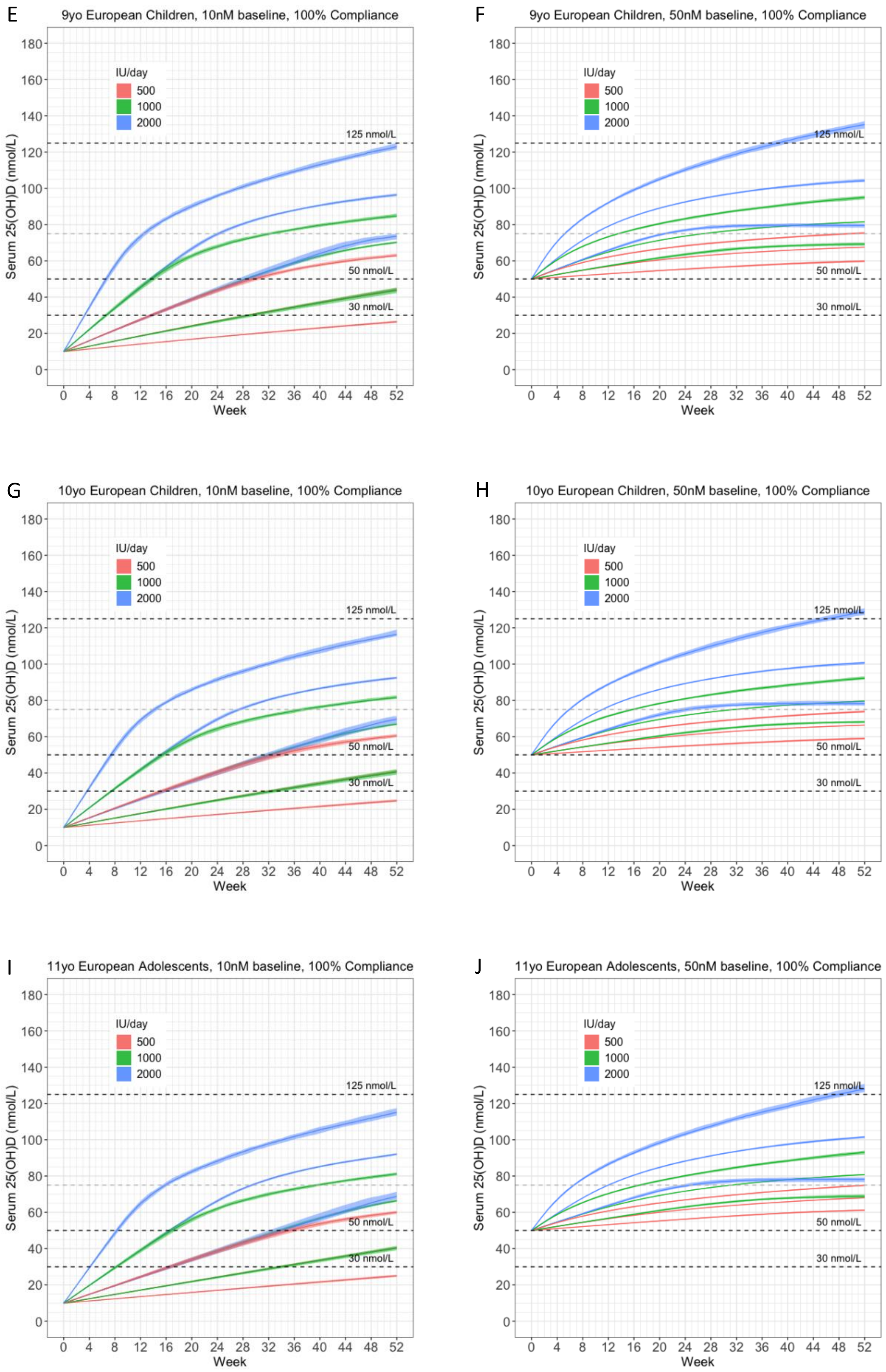
E
9yo European Children, 10nM baseline, 100% Compliance
F
9yo European Children, 50nM baseline, 100% Compliance
G
10yo European Children, 10nM baseline, 100% Compliance
H
10yo European Children, 50nM baseline, 100% Compliance
I
11yo European Adolescents, 10nM baseline, 100% Compliance
J
11yo European Adolescents, 50nM baseline, 100% Compliance
IU/day
500
1000
2000
125 nmol/L
50 nmol/L
30 nmol/L
Serum 25(OH)D (nmol/L)
Week
0
20
40
60
80
100
120
140
160
180
0 4 8 12 16 20 24 28 32 36 40 44 48 52

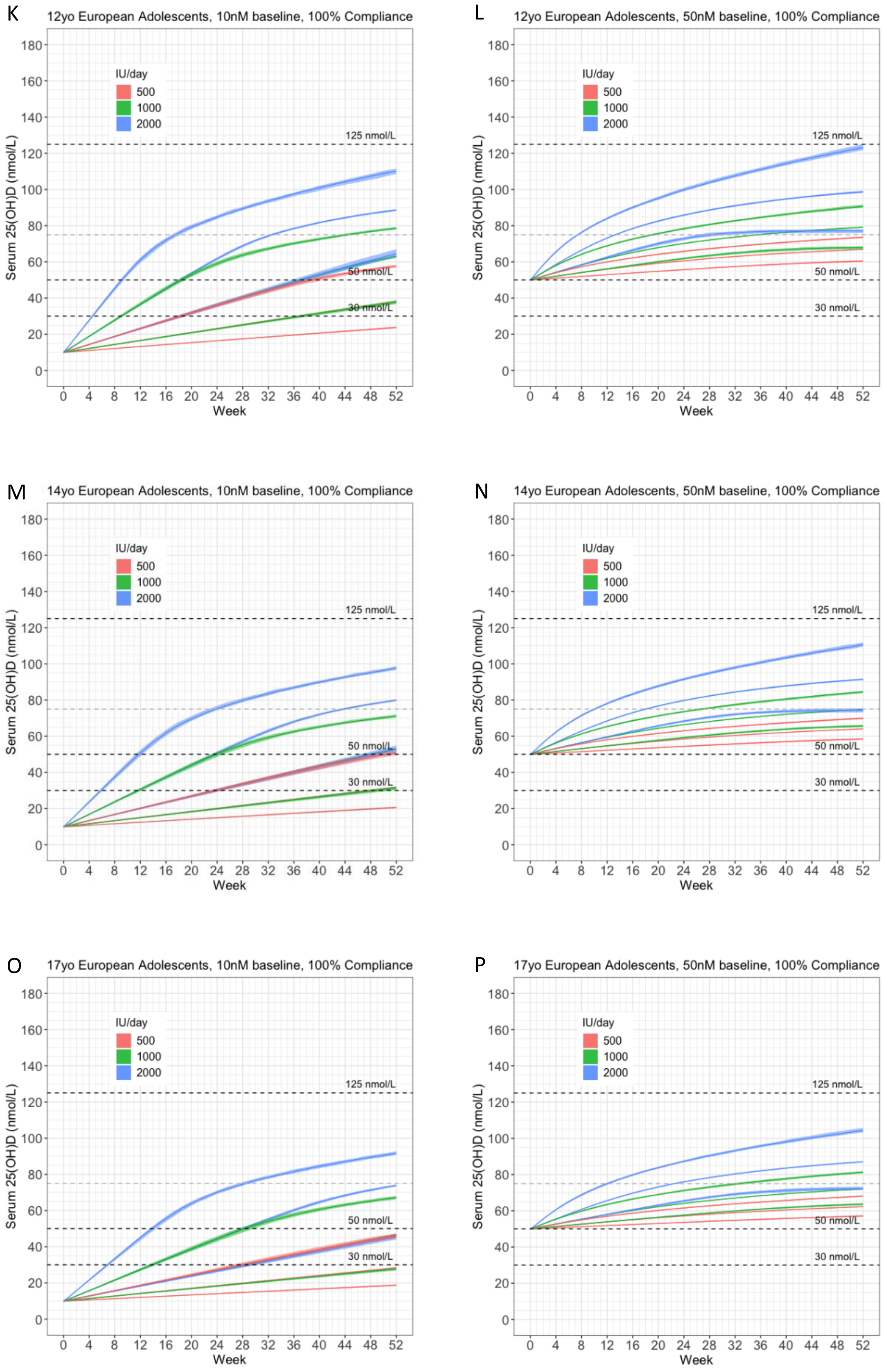
K
12yo European Adolescents, 10nM baseline, 100% Compliance
L
12yo European Adolescents, 50nM baseline, 100% Compliance
M
14yo European Adolescents, 10nM baseline, 100% Compliance
N
14yo European Adolescents, 50nM baseline, 100% Compliance
O
17yo European Adolescents, 10nM baseline, 100% Compliance
P
17yo European Adolescents, 50nM baseline, 100% Compliance
IU/day
500
1000
2000
125 nmol/L
50 nmol/L
30 nmol/L
Serum 25(OH)D (nmol/L)
Week
0 20 40 60 80 100 120 140 160 180
0 4 8 12 16 20 24 28 32 36 40 44 48 52
